# Associations between area-level socioeconomic disadvantage and COVID-19 disease consequences in Sydney, Australia: A retrospective cohort analysis

**DOI:** 10.1101/2025.09.25.25336675

**Authors:** Christopher Standen, Esther Tordjmann, James G Wood, Fiona Haigh

## Abstract

**Background:** Socioeconomic disparities have shaped COVID-19 outcomes worldwide. Focusing on *disease consequences once infected* (severity among cases), we examined whether area-level socioeconomic disadvantage was associated with hospitalisation and death among COVID-19 cases in Greater Sydney, Australia.

**Methods:** We conducted a retrospective cohort study of confirmed and probable COVID-19 cases recorded in the New South Wales Notifiable Conditions Information Management System from 2 March 2020 to 21 February 2022. Area-level disadvantage was measured using the Index of Relative Socio-Economic Disadvantage (IRSD). We modelled the odds of (a) hospitalisation and (b) death conditional on infection using logistic regression, adjusting for age and gender.

**Results:** Among 782,883 included cases, 3.5% were hospitalised and 0.2% died due to COVID-19. Greater area-level disadvantage (lower IRSD) was associated with higher odds of hospitalisation (adjusted odds ratio [AOR] 0.996 per IRSD point; 95% CI 0.996–0.996) and death (AOR 0.997; 95% CI 0.996–0.997), holding age and gender constant. For illustration, the difference between two Sydney postal areas with markedly different IRSD scores corresponds to several-fold differences in the odds of hospitalisation and death among cases.

**Conclusions:** Area-level socioeconomic disadvantage was associated with higher risks of hospitalisation and death among COVID-19 cases in Greater Sydney – a setting with public hospital care – indicating inequities in disease consequences once infected. Given the absence of individual-level comorbidity and vaccination data, the most plausible explanation is disparities in comorbidity and risk-factor burden, although contributions from differences in access to and quality of care cannot be ruled out. Public health responses should prioritise chronic-disease prevention and management in disadvantaged communities to mitigate inequitable outcomes in future pandemics.

## 1 Background

Disparities in COVID-19 disease outcomes have highlighted the role of social determinants of health. While individual risk factors such as older age, sex, and smoking are consistently associated with increased risks of hospitalisation and death [1–3], international evidence shows these risks are amplified among socioeconomically disadvantaged populations. People living in lower-income or deprived areas have experienced higher hospitalisation and mortality risk in the United States, the United Kingdom, and mainland Europe [4,5].

Following Katikireddi et al. [6], who describe six pathways by which social stratification can produce unequal pandemic outcomes, we focus here on those that affect severity among cases: (3) differential disease consequences once infected (our primary target) and, to a lesser extent, (5) differential effectiveness of control measures (e.g., vaccination). The other pathways – (1) differential exposure, (2) differential vulnerability to infection, (4) differential social consequences, and (6) differential consequences of control measures – are outside our analytical scope.

Most studies have focused on disparities in absolute risks of hospitalisation and death. Fewer have examined outcomes conditional on infection – i.e., differential disease consequences (Katikireddi et al.’s third pathway) – which may reflect differences in comorbidity burden and vaccination status, as well as access to and quality of treatment [7].

The Australian context offers an opportunity to investigate differential disease consequences across socioeconomic strata within a health system that provides public hospital care. A cohort study in Western Sydney Local Health District (WSLHD), covering a population of approximately 1.1 million, reported no statistically significant association between area-level socioeconomic disadvantage and a composite measure of severe outcome (death, ICU admission, mechanical ventilation) after adjustment for age, gender, number of comorbidities, and vaccination status, attributing this finding to equitable access to high-quality care [8]. However, that study was restricted to a single health district with relatively limited variation in area-level socioeconomic disadvantage.

This study extends the analysis to the entire Greater Sydney metropolitan area (>5 million residents), which has substantial socioeconomic heterogeneity. Guided by Katikireddi et al.’s framework, we focus on differential disease consequences, examining whether, among reported COVID-19 cases, area-level socioeconomic disadvantage is associated with (a) hospitalisation risk and (b) fatality risk after controlling for age and gender. By moving from a single-district to a metropolitan scale, and by disaggregating adverse outcomes, we aim to provide a clearer picture of equity in COVID-19 outcomes in the context of a health system providing public hospital care.

## 2 Methods

### 2.1 Study design and setting

We conducted a retrospective cohort study to investigate the association between area-level socioeconomic disadvantage and COVID-19 hospitalisation and fatality risks among confirmed and probable cases using routinely collected surveillance data. The study was conducted in Greater Sydney, Australia’s most populous metropolitan region, with an estimated population of 5.2 million in 2021. Area-level socioeconomic disadvantage, as measured using the Index of Relative Socio-Economic Disadvantage (IRSD), varies significantly across the region (Figure 1).

**Figure 1.**
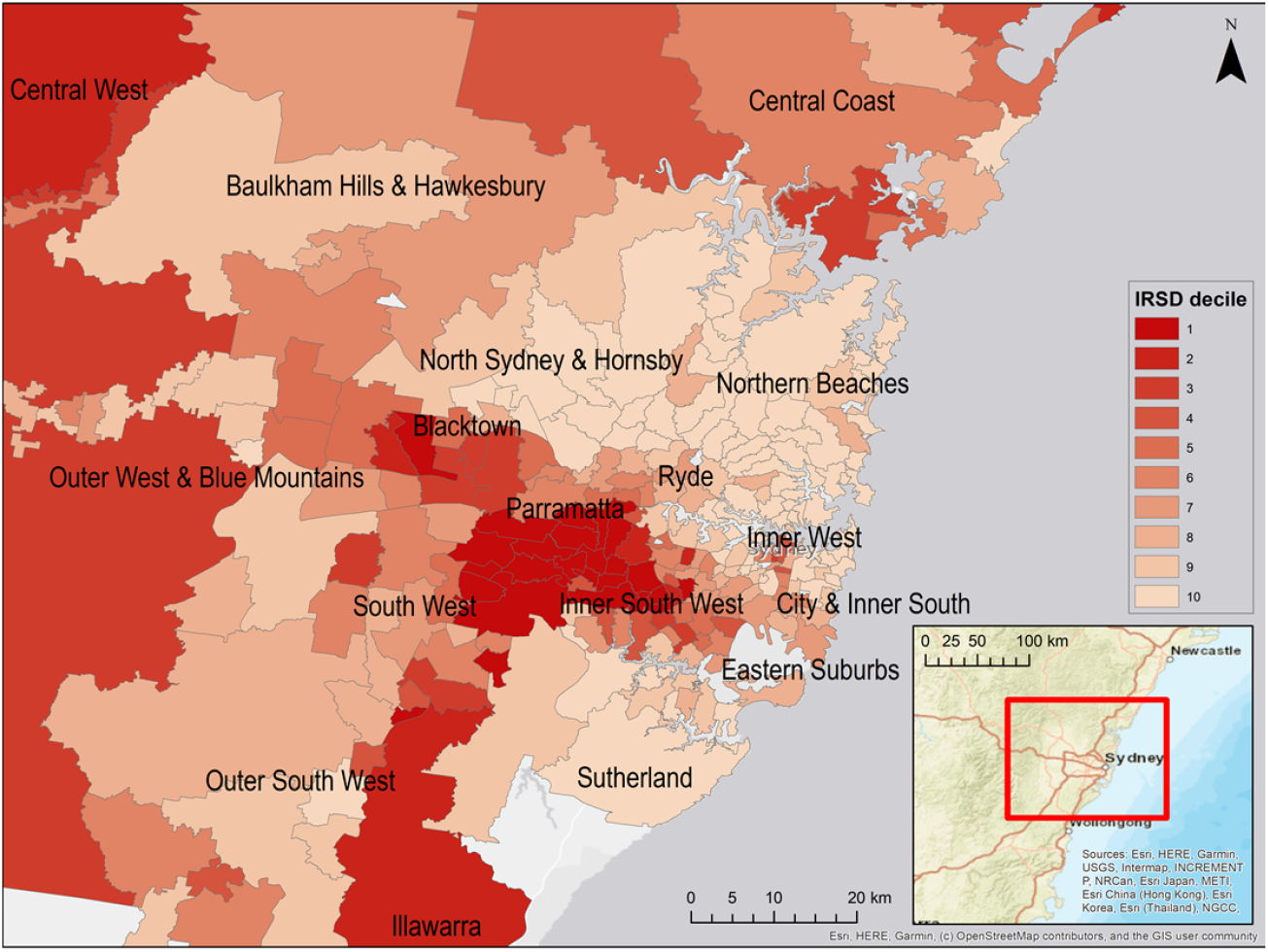
Map of Greater Sydney showing Index of Relative Socio-Economic Disadvantage (IRSD) deciles for postal areas.

The study period commenced on 2 March 2020, when the first locally acquired case of COVID-19 in Greater Sydney was reported, and ended on 21 February 2022, by which time emergency public health restrictions had ended, and the majority of the state’s population had been fully vaccinated [9].

Throughout the study period, polymerase chain reaction (PCR) testing for COVID-19 was widely accessible and free of charge without the need for a referral. Hospital care for COVID-19 was also provided free of charge in public hospitals to all Medicare card holders (Australian citizens, permanent residents, and eligible visitors). Nevertheless, potential barriers to accessing testing and healthcare may have persisted, particularly in socioeconomically disadvantaged communities [7,10].

Efforts were made during the study period to protect older adults and other high-risk populations through infection control measures, particularly in high-risk settings such as aged care facilities [7,11].

This study formed part of a broader, equity-focused health impact assessment (EFHIA) of the COVID-19 pandemic and associated public health responses in Sydney, Australia [7,12].

### 2.2 Data sources

COVID-19 case data were obtained from the Notifiable Conditions Information Management System (NCIMS), maintained by the New South Wales (NSW) Ministry of Health, on 22 February 2022. NCIMS data were de-identified before analysis by the corresponding/lead author (CS). Other authors did not have access to information that could identify individual cases during or after data collection.

Demographic and socioeconomic data were sourced from the 2021 Census of Population and Housing [13] and the Socio-Economic Indexes for Areas (SEIFA) [14], both published by the Australian Bureau of Statistics.

### 2.3 Variables

The primary outcomes were hospitalisation and death due to COVID-19, recorded as dichotomous variables in the NCIMS database. The key independent variable was area-level socioeconomic disadvantage, specifically, the IRSD score corresponding to each case’s residential postal area. IRSD scores were linked algorithmically to NCIMS records. Covariates included age group (categorical) and gender (categorical), also derived from NCIMS records.

### 2.4 Analysis

Initial data cleaning steps involved the exclusion of cases lacking a valid postal area, and cases acquired overseas. Descriptive analysis compared distributions of cases, hospitalisations and fatalities across gender, Indigenous status, and age groups with those of the Greater Sydney population [13]. Descriptive spatial analyses were conducted using ArcGIS software [15] to map incidence of COVID-19 cases, case hospitalisation rate, and case fatality rate. Further exploratory analysis involved examining relationships between area-level socioeconomic disadvantage (IRSD score) and COVID-19 cases, hospitalisations and fatalities using scatterplots and Pearson correlation tests. These analyses were conducted in Python [16].

Multivariate logistic regression models were then applied to assess the associations between IRSD score and two key outcomes: the risk of hospitalisation and the risk of death due to COVID-19. In both models, covariates included age and gender. The Box-Tidwell procedure was used to confirm a linear relationship between IRSD score and the logit transformation of the dependent variable. Variance inflation factors were calculated to confirm the absence of multicollinearity among the independent variables. Regression modelling was carried out using IBM SPSS Statistics [17]. We followed STROBE reporting guidelines for observational studies.

## 3 Results

There were 782,883 reported confirmed or probable COVID-19 cases with a valid postal area in the study area and not acquired overseas during the study period. Of these cases, 27,122 (3.5%) were hospitalised, 2,280 (0.3%) were admitted to an intensive care unit (ICU), and 1,491 (0.2%) were recorded as dying due to COVID-19. The distribution of these cases across gender, age and Indigenous status is presented in Table 1.

**Table 1.** Characteristics of reported COVID-19 cases, hospitalisations and fatalities in Greater Sydney (to 21 February 2022)

| Characteristic | Number of reported COVID-19 cases (N = 782,883) | Number of reported COVID-19 hospitalisations (N = 27,122) | Number of reported COVID-19 fatalities (N = 1,491) | Greater Sydney population 2021 (N = 5,231,147) |
| --- | --- | --- | --- | --- |
| **Gender/sex<sup>a</sup>** |  |  |  |  |
| Female | 393,253 (50.2%) | 14,072 (51.9%) | 580 (38.9%) | 50.6% |
| Male | 388,668 (49.6%) | 13,033 (48.1%) | 911 (61.1%) | 49.4% |
| Transgender | 2 (<0.1%) | 1 (<0.1%) | 0 (0.0%) | - |
| Not stated/inadequately described | 960 (0.1%) | 16 (<0.1%) | 0 (0.0%) | - |
| **Indigenous status** |  |  |  |  |
| Not Indigenous | 257,338 (32.9%) | 17,639 (65.0%) | 1,308 (87.7%) | 94% |
| Indigenous | 18,599 (2.4%) | 964 (3.6%) | 18 (1.2%) | 1.7% |
| Not stated/unknown | 506,946 (64.8%) | 8,519 (31.4%) | 165 (11.1%) | 4.3% |
| **Age group** |  |  |  |  |
| 0–4 | 38,353 (4.9%) | 1,243 (4.6%) | 1 (0.1%) | 6.0% |
| 5–19 | 156,759 (20.0%) | 1,894 (7.0%) | 0 (0.0%) | 18.1% |
| 20–39 | 325,116 (41.5%) | 7,916 (29.2%) | 26 (1.7%) | 30.0% |
| 40–59 | 182,877 (23.4%) | 6,502 (24.0%) | 127 (8.5%) | 25.5% |
| 60+ | 79,709 (10.2%) | 9,567 (35.3%) | 1,337 (89.7%) | 20.5% |
| Not stated/unknown | 69 (<0.1%) | 0 (0.0%) | 0 (0.0%) | - |

<sup>a</sup> For COVID-19 cases, hospitalisations and deaths, this table reports the gender distribution. For the Greater Sydney population, this table reports the sex distribution from the 2021 Census of Population and Housing. The NCIMS does not record sex and does not allow any gender categories other than the four listed to be recorded. The 2021 Census did not allow respondents to report their gender. It allowed respondents to select from three categories for the sex question: male, female, and non-binary sex. However, Census products report data from the sex question as male and female only.

The proportion of cases in the 60+ age group (10.2%) was half the corresponding population proportion (20.5%), which could be partly attributed to enhanced infection control measures for older adults [11].

Figure 2, Figure 3 and Figure 4 show the spatial distributions of cases per 100,000 persons, hospitalisations per 100,000 cases, and fatalities per 100,000 cases, respectively.

**Figure 2.**
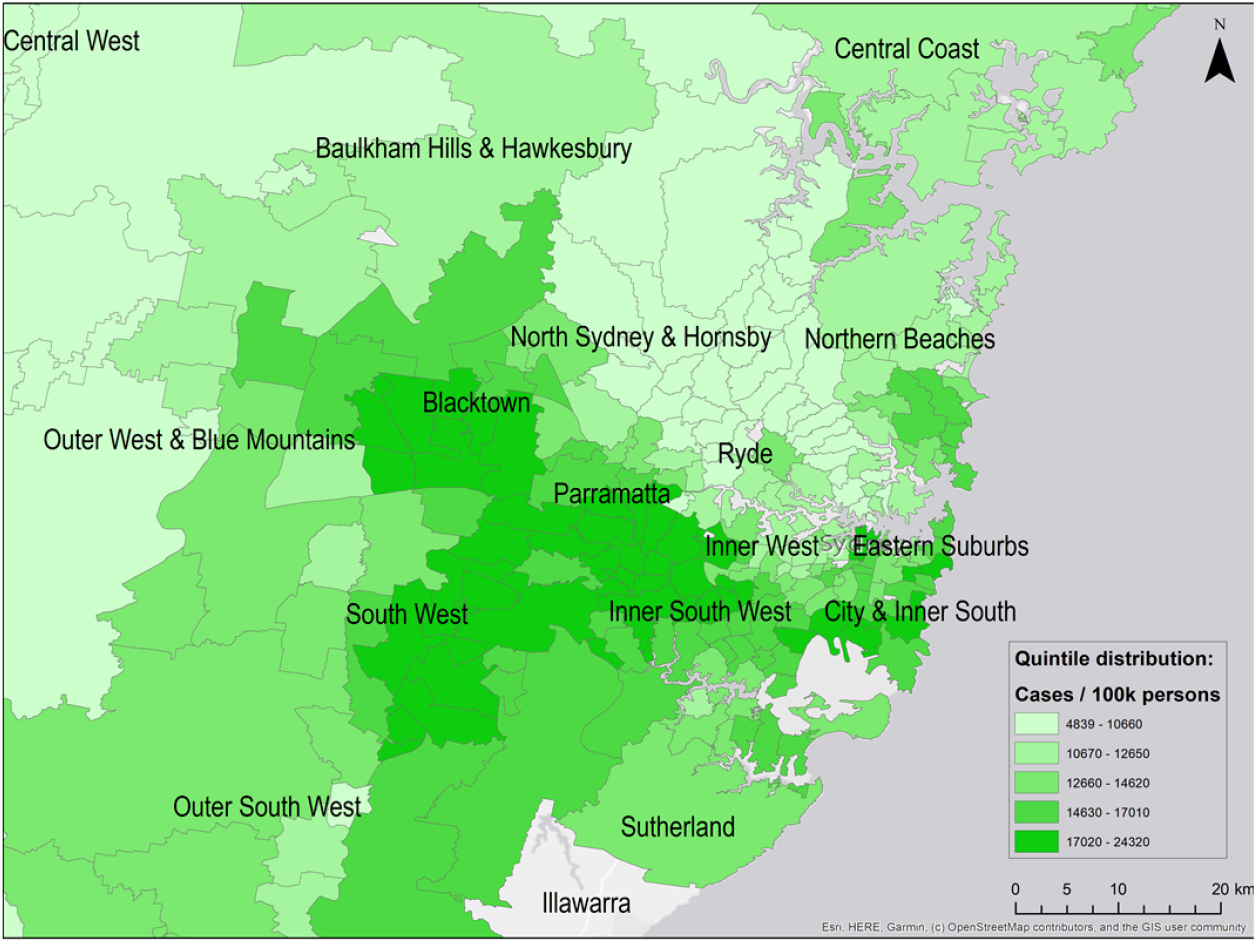
Spatial distribution of reported COVID-19 cases per 100,000 persons across Greater Sydney postal areas (to 21 February 2022)

**Figure 3.**
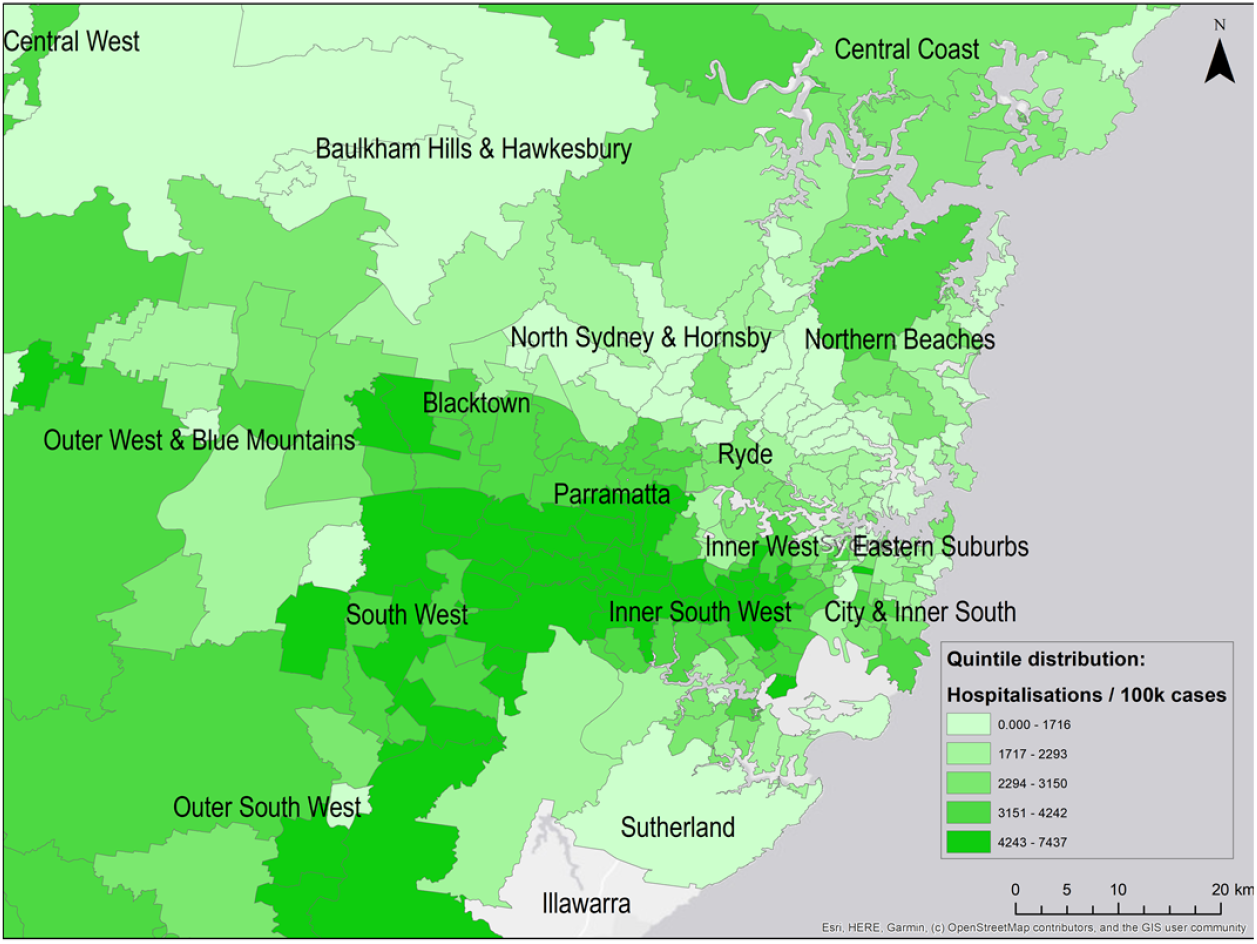
Spatial distribution of reported COVID-19 hospitalisations per 100,000 cases across Greater Sydney postal areas (to 21 February 2022)

**Figure 4.**
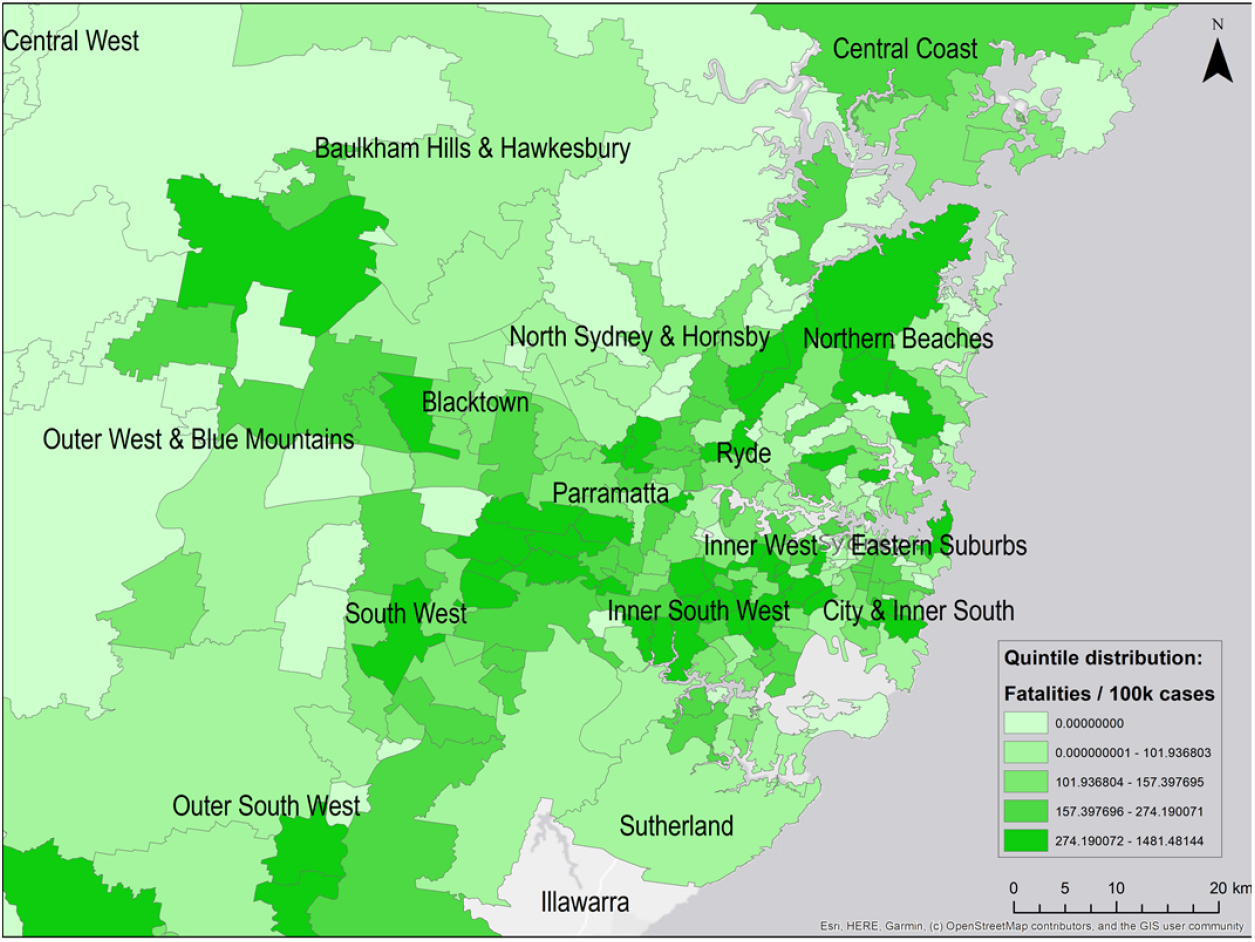
Spatial distribution of reported COVID-19 fatalities per 100,000 cases across Greater Sydney postal areas (to 21 February 2022)

The scatterplots (Figure 5) indicated a linear relationship between IRSD score and COVID-19 outcomes. At the postal area level, there was a strong negative correlation between IRSD score and both case incidence (Figure 5(a)) and hospitalisation risk (Figure 5(b)). There was a weak negative correlation between IRSD score and fatality risk (Figure 5(c)).

**Figure 5.**
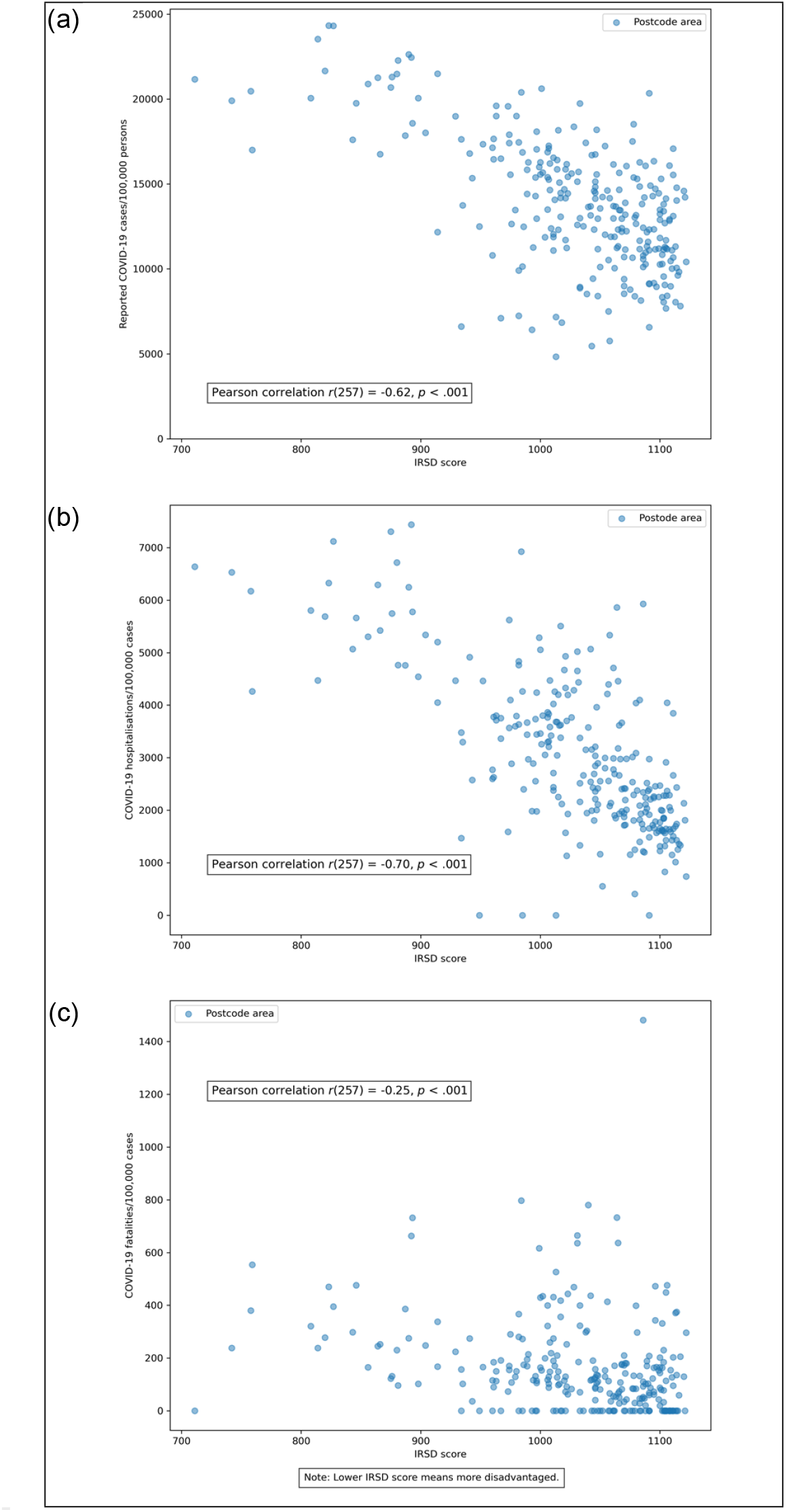
Relationship between area-level socioeconomic disadvantage and (a) reported COVID-19 case incidence, (b) COVID-19 case hospitalisation rate, and (c) COVID-19 case fatality rate.

For the logistic regression analyses of hospitalisation risk and fatality risk, cases with incomplete/unknown information, or belonging to a category with a very low frequency were dropped, leaving 779,774 cases.

The logistic regression model of case hospitalisation risk (Table 2) was statistically significant (*χ*^*2*^(5) = 20,160.19, *p* <.001). The model explained 9.8% (Nagelkerke *R*^*2*^) of the variance in hospitalisation risk and correctly classified 96.5% of cases. Increasing IRSD score (i.e., decreasing socioeconomic disadvantage) was associated with a decrease in the risk of hospitalisation (adjusted odds ratio (AOR) 0.996, 95% CI 0.996–0.996): a case living in Fairfield (postal area 2165; IRSD score 758) was four times as likely to be hospitalised as a case living in Northbridge (postal area 2063; IRSD score 1,122), after adjusting for age and gender differences. Risk of hospitalisation increased with age, with a case aged 50–69 more than three times as likely to be hospitalised as a case aged 0–19 (AOR 3.25, 95% CI 3.11– 3.40), and a case aged 70+ more than 14 times as likely to be hospitalised (AOR 14.59, 95% CI 13.95–15.26). The hospitalisation risk for men was about 8% less than that for women (AOR 0.92, 95% CI 0.90–0.94).

**Table 2.**
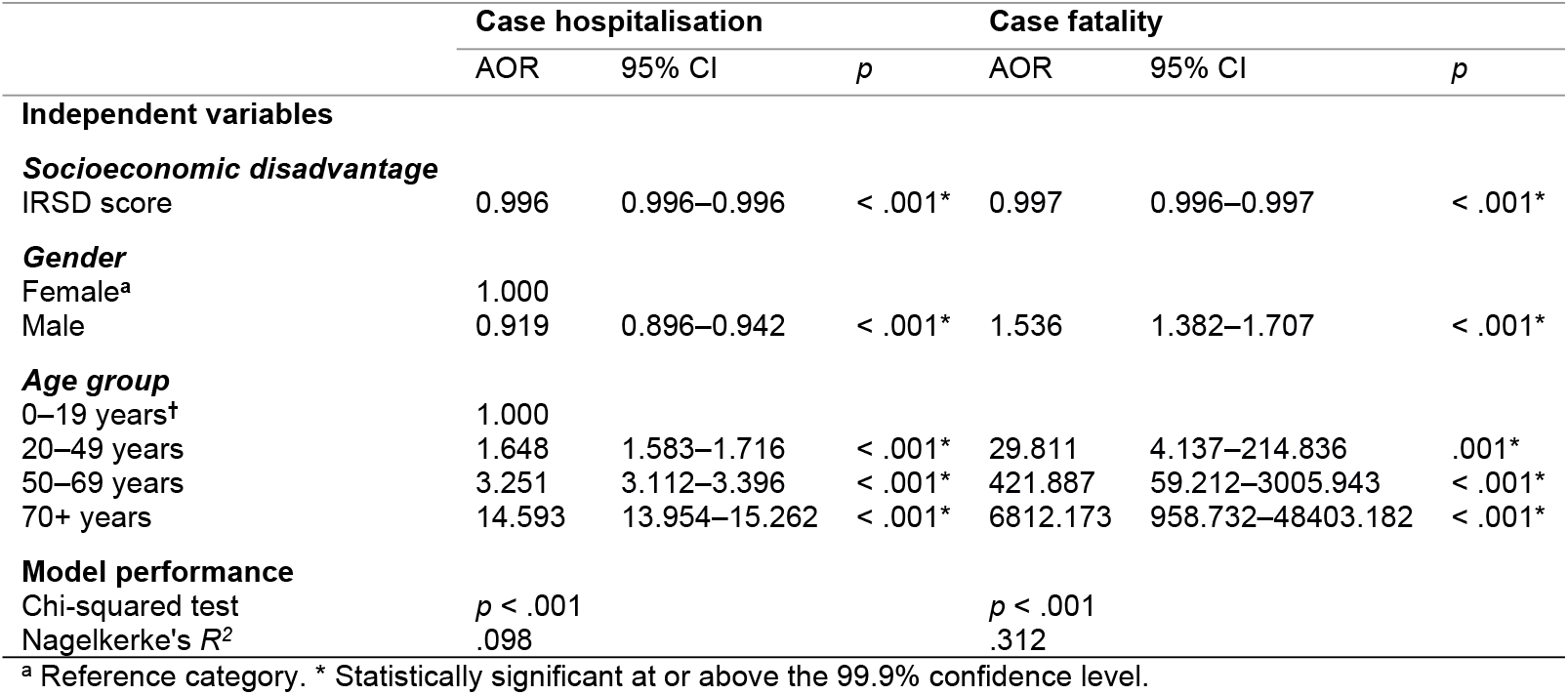
Logistic regression models of COVID-19 case hospitalisation and case fatality.

|  | Case hospitalisation |  |  | Case fatality |  |  |
| --- | --- | --- | --- | --- | --- | --- |
|  | AOR | 95% CI | *p* | AOR | 95% CI | *p* |
| **Independent variables** |  |  |  |  |  |  |
| **Socioeconomic disadvantage** |  |  |  |  |  |  |
| IRSD score | 0.996 | 0.996–0.996 | < .001* | 0.997 | 0.996–0.997 | < .001* |
| **Gender** |  |  |  |  |  |  |
| Female<sup>a</sup> | 1.000 |  |  |  |  |  |
| Male | 0.919 | 0.896–0.942 | < .001* | 1.536 | 1.382–1.707 | < .001* |
| **Age group** |  |  |  |  |  |  |
| 0–19 years<sup>†</sup> | 1.000 |  |  |  |  |  |
| 20–49 years | 1.648 | 1.583–1.716 | < .001* | 29.811 | 4.137–214.836 | .001* |
| 50–69 years | 3.251 | 3.112–3.396 | < .001* | 421.887 | 59.212–3005.943 | < .001* |
| 70+ years | 14.593 | 13.954–15.262 | < .001* | 6812.173 | 958.732–48403.182 | < .001* |
| **Model performance** |  |  |  |  |  |  |
| Chi-squared test | *p* < .001 |  |  | *p* < .001 |  |  |
| Nagelkerke's *R*<sup>2</sup> | .098 |  |  | .312 |  |  |

<sup>a</sup> Reference category. \* Statistically significant at or above the 99.9% confidence level.

Similarly, the logistic regression model of case fatality risk (Table 2) was statistically significant (*χ*^*2*^(5) = 6,684.97, *p* <.001). The model explained 31.2% (Nagelkerke *R*^*2*^) of the variance in fatality risk and correctly classified 99.8% of cases. Increasing IRSD score (i.e., decreasing socioeconomic disadvantage) was associated with a decrease in the risk of death due to COVID-19 (AOR 0.997, 95% CI 0.996–0.997): a case living in Fairfield (postal area 2165; IRSD score 758) was three times as likely to die due to COVID-19 as a case living in Northbridge (postal area 2063; IRSD score 1,122), after adjusting for age and gender differences. Risk of death due to COVID-19 increased with age: a case aged 50–69 was more than 400 times as likely to die as a case aged 0–19 (AOR 421.89, 95% CI 59.21– 3,005.94) and a case aged 70+ was more than 6,000 times as likely to die due to COVID-19 (AOR 6812.17, 95% CI 958.73–48,403.18). Male cases were more likely to die due to COVID-19 than female ones (AOR 1.54, 95% CI 1.38–1.71).

## 4 Discussion

This study investigated associations between area-level socioeconomic disadvantage and disease consequences once infected with SARS-CoV-2 – the third pathway in Katikireddi et al.’s framework for understanding COVID-19 inequities [6] – in the Greater Sydney metropolitan region. After controlling for age and gender, COVID-19 cases residing in more socioeconomically disadvantaged postal areas had higher odds of hospitalisation and of death due to COVID-19. These results indicate that area-level socioeconomic disadvantage is associated with more adverse COVID-19 consequences conditional on infection.

Male cases were less likely than female cases to be hospitalised but more likely to die due to COVID-19. This may reflect biological differences and differences in admission thresholds (e.g., pregnancy-related admissions among women).

Our findings differ from a cohort analysis confined to Western Sydney Local Health District, which reported no association between area-level socioeconomic disadvantage and a composite measure of severe outcomes, after adjustment for age and comorbidities [8].

Differences in population size, socioeconomic heterogeneity, outcome definition, inclusion of deaths occurring outside hospital, model specification, and ability to adjust for comorbidities and vaccination status may account for the contrasting findings.

While our study design does not allow for the causes of inequalities in disease consequences to be determined, evidence from elsewhere suggests that disparities in comorbidity burden, smoking history, and in access to and quality of healthcare, may have played a role.

Socioeconomic disadvantage is associated with higher prevalence of risk factors for severe COVID-19 – such as smoking, cardiovascular disease, diabetes, obesity, and chronic respiratory disease [1–3]. The WSLHD study’s null association after comorbidity adjustment is consistent with this mechanism, though it did not report if severe outcomes were associated with socioeconomic disadvantage when the comorbidities covariate was omitted from the model.

Despite Australia’s public hospital system, disparities in access to and quality of healthcare, cannot be ruled out as a contributory factor. The broader equity-focused health impact assessment (EFHIA) of the COVID-19 pandemic and associated public health responses in Sydney [7,12] – of which this study forms one component – documented healthcare access barriers during the pandemic (e.g., longer wait times to see general practitioners in more disadvantaged areas, and exclusion from telehealth services for people without Internet access or low digital literacy). The EFHIA also found that infection control measures (e.g., limits on gatherings) disrupted community networks, informal support systems and culturally appropriate community health services.

### 4.1 Strengths and limitations

Strengths include metropolitan-wide coverage, substantial socioeconomic heterogeneity, and the inclusion of deaths in non-hospital settings (e.g., aged care). The analysis explicitly targets disease consequences once infected.

Key limitations include the use of an aggregate, area-level measure of socioeconomic disadvantage (IRSD) to infer the socioeconomic status of individual cases. Furthermore, the IRSD index is an imperfect measure of socioeconomic disadvantage and includes some potentially problematic variables [18], such as the percentage of households with no motor vehicle (the local government area with the lowest rate of household motor vehicle ownership, City of Sydney, is one of the least disadvantaged in Greater Sydney). We were unable to adjust for potentially important covariates, including vaccination status, comorbidities, and smoking history, due to limitations in the available data. This is reflected in the relatively low goodness-of-fit values in the regression models. Indigenous status was poorly recorded in the NCIMS database, with 65% of records being “not stated/unknown”, which meant we were unable to adjust for this variable. Our study spans multiple pandemic phases (variants, vaccination rollout, evolving clinical practice), and we did not adjust for calendar time/wave; thus, part of the observed socioeconomic gradient may reflect time-varying differences across areas rather than inequities in disease severity per se. Finally, the observational design precludes causal attribution of specific mechanisms of adverse COVID-19 disease consequences.

### 4.2 Implications

Despite the above limitations, the findings underscore the importance of recognising and addressing systemic inequities in health and the social determinants of health in pandemic preparedness and response. This includes intensifying chronic disease prevention and management in disadvantaged communities, addressing direct and indirect healthcare access barriers (including those created through pandemic control measures) [10], and investing in equity infrastructures – for example, place-based services and community partnerships that can be rapidly activated during emergencies [19]. Aside from potentially reducing disparities in disease consequences once infected, these measures may also temper other pathways through which disadvantage can lead to higher risk of hospitalisation and death, i.e., differential exposure, differential susceptibility to infection, and differential effectiveness of control measures [6].

The study also highlights the need for good-quality surveillance data, in particular, accurate and consistent recording of Indigenous status, and the ability to link them with clinical and vaccination data at metropolitan, state or national scale.

## 5 Conclusions

This study found that individuals residing in more socioeconomically disadvantaged areas of Greater Sydney were more likely to be hospitalised and more likely to die due to COVID-19 once infected with SARS-CoV-2, even within the context of a public healthcare system.

Reducing inequities in the burden of future health emergencies will require measures that address differentials in exposure, susceptibility to infection, disease severity, and effectiveness of control measures across socioeconomic strata.

## List of abbreviations

AOR: Adjusted odds ratio
EFHIA: Equity-focused health impact assessment ICU Intensive care unit
IRSD: Index of Relative Socio-Economic Disadvantage
NCIMS: Notifiable Conditions Information Management System NSW New South Wales
PCR: Polymerase chain reaction
SARS-CoV-2: Severe acute respiratory syndrome coronavirus 2
SEIFA: Socio-Economic Indexes for Areas
STROBE: Strengthening the Reporting of Observational Studies in Epidemiology

## Declarations

### Ethics approval and consent to participate

Ethics approval for the study was granted by the Sydney Local Health District Research Ethics and Governance Office on 11 December 2020 (Protocol No. X20-0467 and 2020/ETH02564).

### Availability of data and materials

The data underlying this study are third-party public health surveillance records held by NSW Health and cannot be publicly shared due to legal and ethical restrictions under NSW Health data governance and privacy policies. Qualified researchers may request access to NCIMS data through NSW Health’s data governance processes (contact: or the NSW Health data-sharing portal). Access is granted to researchers who meet criteria for confidential data access and have approved ethics and governance documentation.

### Authors’ contributions

CS: Conceptualisation, Methodology, Formal analysis, Investigation, Data curation, Writing – original draft, Visualisation, Project administration. ET: Conceptualisation, Writing – review & editing. JW: Writing – review & editing. FH: Writing – review & editing, Supervision, Project administration, Funding acquisition.

